# Household transmission of SARS-CoV-2 R.1 lineage with spike E484K mutation in Japan

**DOI:** 10.1101/2021.03.16.21253248

**Authors:** Yosuke Hirotsu, Masao Omata

## Abstract

We aimed to investigate SARS-CoV-2 emerging lineage harboring variants in receptor binding domain (RBD) of spike protein in Japan. Total nucleic acids were subjected to whole genome sequencing on samples from 133 patients with coronavirus disease (COVID-19). We obtained the SARS-CoV-2 genome sequence from these patients and examined variants in RBD. As a result, three patients were infected with SARS-CoV-2 harboring E484K mutation in January 2021. These three patients were relatives; one was in the 40s, and two were younger than 10 years old. They had no history of staying abroad and were living in Japan. This strains were classified into GR clade (GISAID), 20B clade (Nextstrain) and R.1 lineage (PANGO). As of March 5, 2021, the R.1 lineage have been identified in 305 samples and dominantly observed in the USA (44%, 135 / 305) and Japan (28%, 84 / 305) from the GISAID database. During the period between October 26, 2020 and February 23, 2021, the frequency of the R.1 lineage was 0.97% (84 / 8,629) of the total confirmed data in Japan and 0.15% (135 / 90,450) in the USA. Although SARS-CoV-2 R.1 lineage was not globally predominant as of March 2021, further analysis is needed to determine whether R.1 variant will disappear or expand in the future.

## Introduction

SARS-CoV-2 has the largest genome (29.9 kb in length) among RNA viruses. The virus encodes an exoribonuclease with a 3′-5′ proofreading function, resulting in slow evolution and lower mutation rate than other RNA viruses [1, 2]. Most occurring mutations are likely to neutral during the evolution and not to influence viral properties. However, some of the mutations are selected due to the change of viral fitness, virulence and transmissibility [3, 4]. In fact, several SARS-CoV-2 variants of concern have emerged within the last few months, especially in three major lineages (B.1.1.7, B.1.351, and P.1) [5-7]. These emerging lineages are characterized by multiple mutations in spike proteins, raising concerns that they may escape monoclonal antibody-therapy or vaccine-elicited antibodies. The B.1.1.7 lineage is estimated to have emerged in late September 2020 and rapidly became the dominant strain in the United Kingdom [5, 8, 9]. The B.1.351 lineage rapidly became the dominant strain in South Africa where it was first detected in October 2020 [6]. P.1 lineage was first identified in four travelers from Brazil and has been associated with cases of reinfection [7, 10].

The hallmark mutation of B.1.1.7, B.1.351, and P.1 is N501Y located in the receptor binding domain (RBD) of the spike protein [5]. The N501Y variant is thought to be more transmissible and possibly more virulent [8, 11-14]. The other hallmark mutation of B.1.351, and P.1 is E484K, which is related to reduce the neutralizing activity of antibodies and raising the concerns about vaccine efficacy [15-20].

In this study, we conducted genetic surveillance and identified R.1 lineage harboring E484K mutation in RBD by whole genome sequencing. The R.1 lineage was observed in three patients and transmitted among relatives in Japan.

## Methods

### Patients and sample collection

A total of 166 COVID-19 patients were confirmed in our hospital, from which 133 were selected for subsequent genome analysis. Nasopharyngeal swab samples were collected using cotton swabs and placed in 3 mL of viral transport media (VTM) purchased from Copan Diagnostics (Murrieta, CA, USA). We used 200 μL of VTM for nucleic acid extraction within 2 h of sample collection.

### Viral nucleic acid extraction

Total nucleic acid was isolated from the samples using the MagMAX Viral/Pathogen Nucleic Acid Isolation Kit (Thermo Fisher Scientific, Waltham, MA, USA) on the KingFisher Duo Prime System (Thermo Fisher Scientific) as we previously described [21, 22]. Briefly, we added 200 µL of VTM, 5 µL of proteinase K, 265 μL of binding solution, 10 μL of total nucleic acid-binding beads, 0.5 mL of wash buffer, and 0.5–1 mL of 80% ethanol to each well of a deep-well 96-well plate. Nucleic acids were eluted with 70 μL of elution buffer.

### Quantitative reverse transcription polymerase chain reaction (RT-qPCR)

Following the protocol developed by the National Institute of Infectious Diseases (NIID) in Japan [23], we performed one-step RT-qPCR to detect SARS-CoV-2. This PCR amplifies the *nucleocapsid* (*N*) gene of SARS-CoV-2 (NC_045512.2). For the internal positive control, the human ribonuclease P protein subunit p30 (*RPP30*) gene was used (Integrated DNA Technologies, Coralville, IA, USA). RT-qPCR assays were conducted on a StepOnePlus Real-Time PCR System (Thermo Fisher Scientific) with the following cycling conditions: 50°C for 5 min for reverse transcription, 95°C for 20 s, and 45 cycles of 95°C for 3 s and 60°C for 30 s. The absolute copy number of viral loads was determined using a serial diluted DNA control targeting the *N* gene of SARS-CoV-2 (Integrated DNA Technologies) as previously described [24, 25].

### Whole genome sequencing

SARS-CoV-2 genomic DNA was amplified using the Ion AmpliSeq SARS-CoV-2 Research Panel consisting of two primer pools covering 99.7% of the viral genome (Thermo Fisher Scientific) [26]. Extracted nucleic acids were subjected to genome sequencing on the Ion Torrent Genexus System according to the manufacturer’s instruction. Sequencing reads and quality were processed on Genexus Software with SARS-CoV-2 plugins. The sequencing reads were mapped and aligned on the reference genome of SARS-CoV-2 strain Wuhan-Hu-1 using the torrent mapping alignment program (TMAP). After initial mapping, a variant call was performed using the Torrent Variant Caller. The COVID19AnnotateSnpEff plugin was used for annotation of variants. Assembly was performed with the Iterative Refinement Meta-Assembler (IRMA) [27], which produced the FASTA file.

### The clade and lineage classification

The viral clade and lineage were conducted using the Global Initiative on Sharing Avian Influenza Data (GISAID) database [28], Nextstrain [29] and Phylogenetic Assignment of Named Global Outbreak Lineages (Pangolin; https://cov-lineages.org/index.html) [30]. The sequence of the identified R.1 variant was deposited to the GISAID EpiCoV database (Accession No. EPI_ISL_1164927, EPI_ISL_1164928 and EPI_ISL_1164929).

Global sequencing data of the R.1 variant was exported from the GISAID EpiCoV database on March 5, 2021. We searched 305 available metadata of R.1 lineage between October 26, 2020 and February 23, 2021. We also searched a total number of samples as following parameter: lineage “R.1”, host “human”, location “Asia / Japan” or “North America / USA”, collection date “October 26, 2020 to February 23, 2021”. During this period, a total of 8,629 samples were registered in Japan, and 90,450 samples were registered in the USA.

### Ethical statement

The Institutional Review Board of the Clinical Research and Genome Research Committee at Yamanashi Central Hospital approved this study and the use of an opt-out consent method (Approval No. C2019-30). The requirement for written informed consent was waived owing to the observational study and the urgent need to collect COVID-19 data. Participation in the study by patients was optional. All methods were performed in accordance with the relevant guidelines and regulations and with the Helsinki Declaration.

## Results

### Household transmission of SARS-CoV-2 spike E484K

To conduct the genomic surveillance, we have started to investigate the variants in RBD of spike protein in Kofu city, Japan [26]. The nucleic acids were stored at our hospital were subjected to whole genome analysis.

We previously identified the P.1 lineage harboring K417T/ E484K / N501Y in a patient [26]. Subsequently, consecutive analysis detected spike E484K mutation in three patients. The samples were collected from these patients in January 2021. These patients were relatives of each other; one was in the 40s, and two were younger than 10 years old. They had no history of staying in foreign countries and lived in Japan. Around the same time, another relative was also infected with SARS-CoV-2, but the samples could not be available because the relative was tested at another hospital. These results suggested they were infected with the same virus lineage and household transmission was occurred.

### Genetic characterization was consistent with R.1 lineage

Sequencing analysis identified 21 variants in total, including 13 missense, 6 synonymous and two intergenic variants. There were four missense mutations in spike protein (W152L, E484K, D614G, G769V), four in orf1ab (T4692I, N6301S, L6337M, I6525T), one in membrane protein (F28L) and four in nucleocapsid protein (S187L, R203K, G204R and Q418H).

To examine the phylogeny based on genetic variations, we analyzed genomic data using GISAID, Nextstrain and Pangolin. The sequence was assigned as GR clade (GISAID), 20B clade (Nextstrain) and R.1 lineage (PANGO). R.1 lineage is the sublineage of B.1.1.316 (github.com/cov-lineages/pangolin).

### Epidemiological event of R.1 lineage

To investigate the global distribution of R.1 lineage, we next collected registration data from the EpiCoV of GISAID database [28]. As of March 5, 2021, a total of 305 samples of R.1 lineage had been registered from all over the world, with the majority spread in the USA (44%, 135/305) and Japan (28%, 84/305) (Figure 1A and Table 1). R.1 lineage was first reported in Texas, USA at the end of October 2020, and was found in Japan at the end of November 2020. The number of detected lineages has changed in a similar trend between the USA, Japan and other countries (Figure 1A), implying that SARS-CoV-2 R.1 lineage may have emerged in several regions at approximately the same time.

**Table 1.**
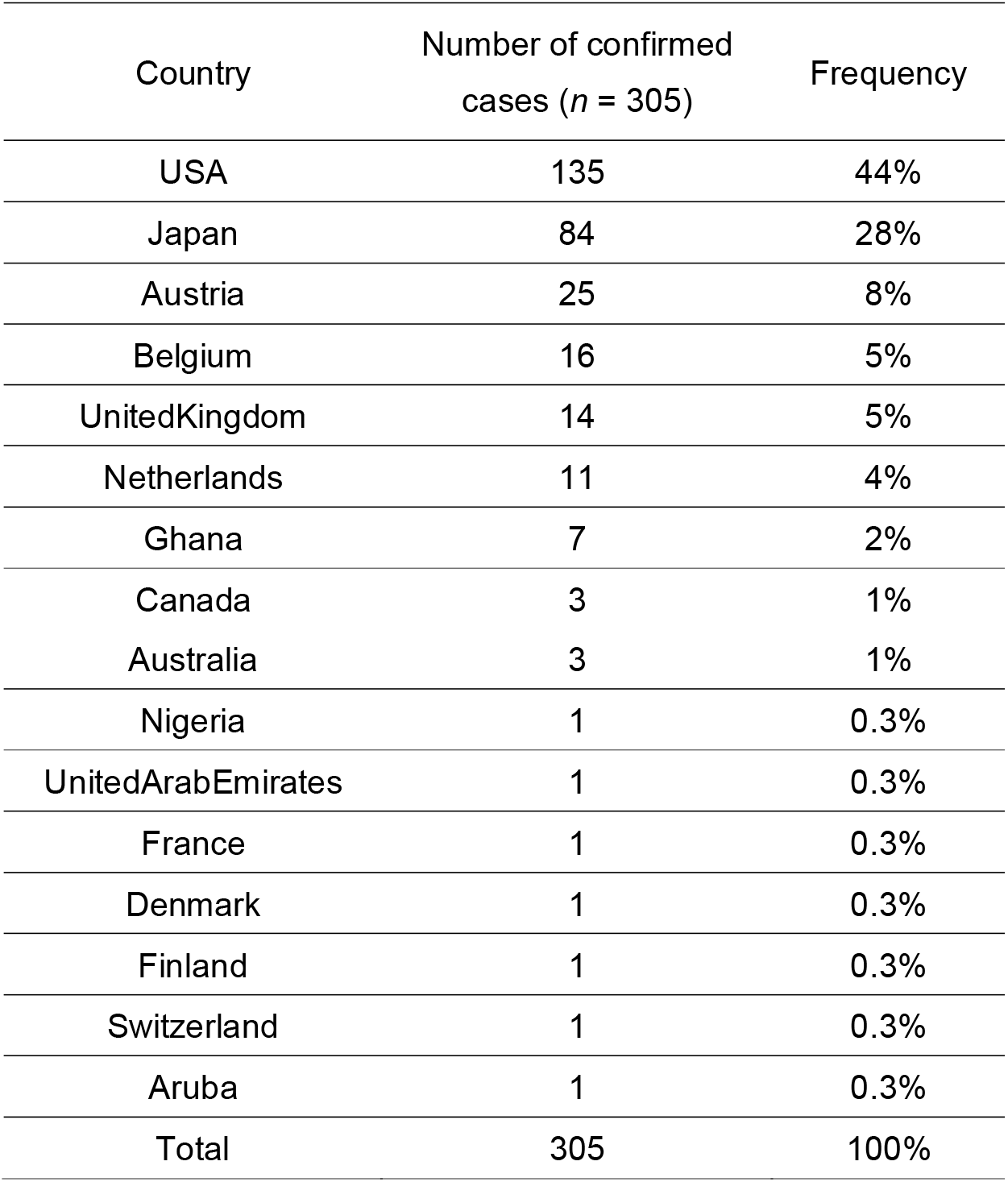
Frequency of R.1 strains across countries.

| Country | Number of confirmed cases (*n* = 305) | Frequency |
| --- | --- | --- |
| USA | 135 | 44% |
| Japan | 84 | 28% |
| Austria | 25 | 8% |
| Belgium | 16 | 5% |
| UnitedKingdom | 14 | 5% |
| Netherlands | 11 | 4% |
| Ghana | 7 | 2% |
| Canada | 3 | 1% |
| Australia | 3 | 1% |
| Nigeria | 1 | 0.3% |
| UnitedArabEmirates | 1 | 0.3% |
| France | 1 | 0.3% |
| Denmark | 1 | 0.3% |
| Finland | 1 | 0.3% |
| Switzerland | 1 | 0.3% |
| Aruba | 1 | 0.3% |
| Total | 305 | 100% |

**Figure 1.**
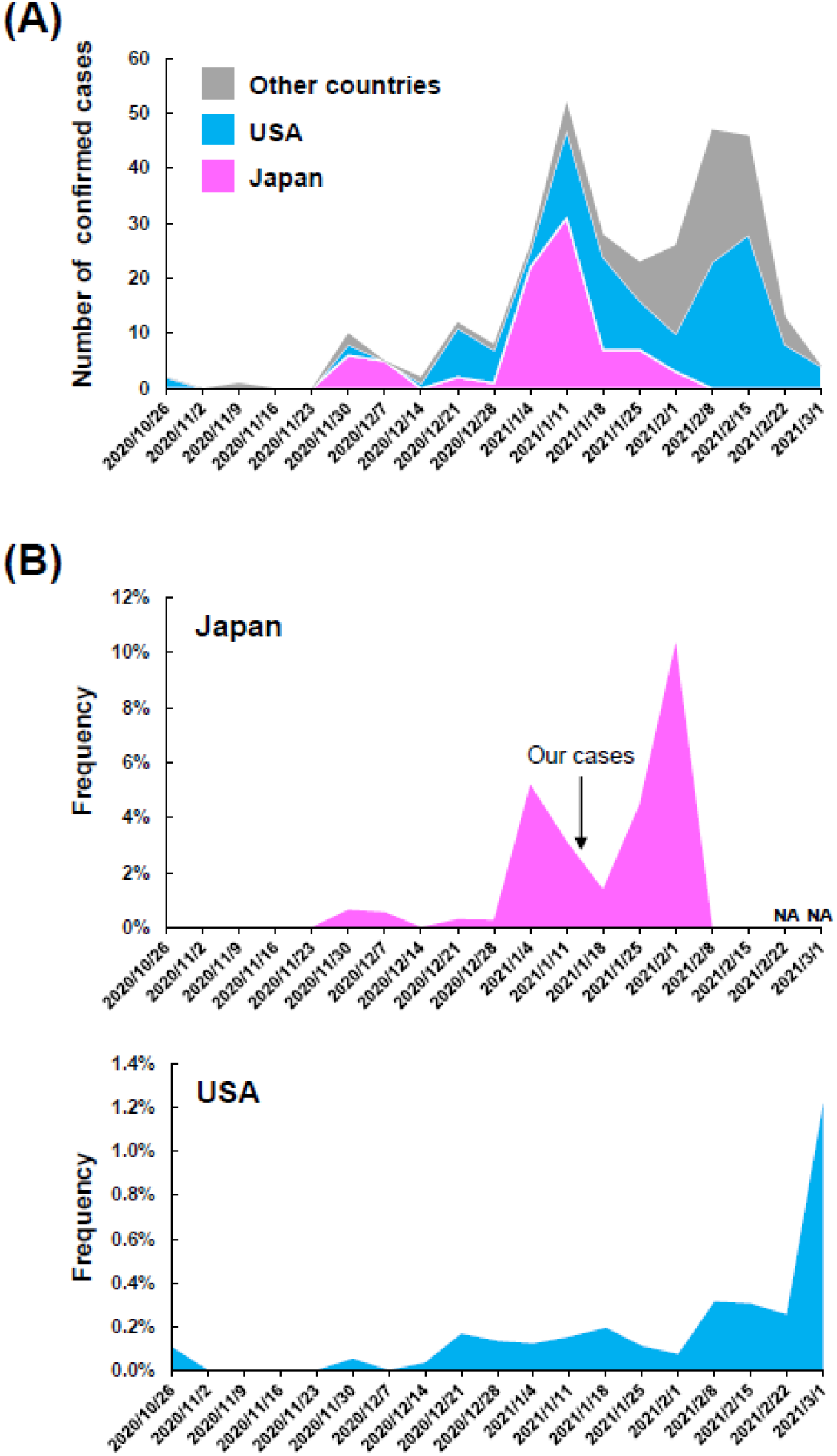
Timeline and frequency of emergence of R.1 lineage in the world. (**A**) Timeline of the number of confirmed cases of R.1 lineage. The plot indicates data for Japan (pink), the USA (light blue), and other countries (gray). This plot was created based on the data in Table 1. (**B**) The frequency of R.1 strains relative to the total number of registered samples during the period is shown for Japan (upper panel) and the USA (lower panel). The arrow indicates the time of infection of the three individuals identified in this study. NA, not applicable.

The overall detection frequency was 0.97% (84 / 8,629) in Japan and 0.15% (135 / 90,450) in the USA over the observed period. The frequency of weekly data showed an increase from mid-January to early February 2021 in Japan and from early February to early March in the USA (Figure 1B). The frequency of the Japanese data is not available because the any data was not registered after 13 February 2021, but it is possible that R.1 lineage is gradually increasing in Japan.

## Discussion

The development and approval of a safe and effective vaccine against SARS-CoV-2 give us a great hope eliminating the virus. Vaccines have been approved in many countries within a year and this event is an unprecedented scientific achievement [31]. On the other hand, mutations in RBD are of great concern because of the potential for immune escape.

Actually, many neutralizing antibodies involving the RBD or the N-terminal domain [32]. Most convalescent sera and mRNA vaccine-elicited sera had reduced neutralizing activity against SARS-CoV-2 containing the E484K spike mutation [18-20]. Mutations in the RBD result in tighter binding of ACE2, allowing it to escape extensive neutralization of monoclonal antibodies mainly by E484K [33].

In Brazil, patients were re-infected with the virus carrying the E484K mutation, which is supported by the evidence that E484K is related to escape neutralizing antibodies in recovered patients. Recently, the E484K mutation present in another SARS-CoV-2 variant of concern (B.1.526) identified in New York [34, 35]. In this study, R.1 lineage harboring E484K mutation was identified in three family members in Japan. The circulating trend of R.1 lineage showed similar pattern in each country, implying that the ancestral strain acquired a homegrown variant almost simultaneously, and subsequently R.1 lineage are likely to emerge.

The SARS-CoV-2 acquired around one or two mutations per month. This seems to be very slow. However, the more the virus circulates in population, the more opportunity it has to change [36]. Almost half of all genome sequences are deposited from the United Kingdom in the GISAID database [31]. However, not enough samples have been sequenced in many countries, therefore, new emerging strain have been missed possibly. Therefore, the identification of mutants by genomic surveillance in real time will provide us with important insights into the effectiveness of vaccines, the development of antibody therapy and public health.

## Data Availability

All data are described in this manuscript.

## Acknowledgments

This study was supported by a Grant-in-Aid for the Genome Research Project from Yamanashi Prefecture (to M.O. and Y.H.), the Japan Society for the Promotion of Science (JSPS) KAKENHI Early-Career Scientists JP18K16292 (to Y.H.), a Grant-in-Aid for Scientific Research (B) 20H03668 (to Y.H.), a Research Grant for Young Scholars (to Y.H.), the YASUDA Medical Foundation (to Y.H.), the Uehara Memorial Foundation (to Y.H.), and Medical Research Grants from the Takeda Science Foundation (to Y.H.). We thank Hitoshi Mochizuki for applying for the ethics committee. We also thank Masato Kondo, Ryota Tanaka, and Kazuo Sakai (Thermo Fisher Scientific) for technical help, all of the medical and ancillary hospital staff, and the patients for their participation.

